# Personalized Brain-Based Analgesia Detection with Portable fNIRS and AI

**DOI:** 10.64898/2026.05.20.26353377

**Authors:** Cristian Minoccheri, Pangyu Joo, Xiao-Su Hu, Hafsa Affendi, Fadi Elayyan, Angeline Harville, Neville J. McDonald, Tatiana Botero, Alexandre F. DaSilva

## Abstract

Neuroimaging-based pain decoding faces two underappreciated challenges: between-subject variability that prevents classifiers from generalizing across patients, and within-session cross-validation designs that inflate reported accuracy by conflating within- and between-person variance. Here we address both using portable functional near-infrared spectroscopy (fNIRS) during pharmacologically verified local nerve anesthesia. Twenty-five patients with clinically painful teeth underwent 36-channel bilateral fNIRS during percussion before ("Pre") and after ("Post") local nerve anesthesia. In 13 block-success patients, a paired Pre-versus-Post comparison with healthy-tooth control identified three temporal hemodynamic response function (HRF) features – late slope, mean first derivative, and baseline-normalized amplitude – whose analgesia interaction effects (d = 0.63–0.79) exceeded that of raw general linear model (GLM) amplitude (d = 0.56), with a significant difference-in-differences interaction (p = 0.011). Per-patient calibration with these features yielded leave-one-subject-out (LOSO) AUC = 0.68–0.76 for nonlinear classifiers (permutation p = 0.002), with HbO-specific feature selection achieving the best performance (RF AUC = 0.760); a healthy-tooth negative control was non-significant. End-to-end deep learning on raw time series (CNN-LSTM AUC = 0.719) was competitive with feature-based classifiers, while linear models did not reach significance. Critically, head-to-head comparison of within-session CV and LOSO on the same data revealed mean inflation of +0.13 AUC across all model types, including deep learning – demonstrating that high within-session accuracy alone does not establish subject-independent validity. Exploratory analyses suggested complementary roles for oxyhemoglobin (HbO; within-patient analgesia detection) and deoxyhemoglobin (HbR; cross-patient information), and that trial-to-trial response variability may complement amplitude for cross-patient pain detection. These results show that per-patient calibration with temporal HRF features supports subject-independent analgesic-state detection under strict LOSO evaluation, and that within-session validation – standard in the fNIRS pain-decoding literature – can substantially overestimate performance.

## 1. Introduction

Decoding pain from brain signals is a central challenge in clinical neuroimaging. Functional magnetic resonance imaging (fMRI) has shown that nociception activates distributed cortical networks (Apkarian et al., 2005; Tracey and Mantyh, 2007) and yielded multivariate pain signatures such as the Neurologic Pain Signature (NPS) (Wager et al., 2013). However, the NPS shows a large within-person effect (d = 1.45) but only a moderate between-person effect (d = 0.49) (Han et al., 2022), and higher-order brain regions show larger individual variability in their pain representations than somatosensory cortex (Kohoutova et al., 2022). A recent systematic review of 57 neuroimaging pain-decoding studies identified sample size, spatial scale, and modeling approach as key determinants of classifier performance, and highlighted the need for rigorous cross-subject validation (Kohoutova et al., 2025). Cross-patient generalization therefore appears limited not by absent signal, but by between-individual scaling – and whether a given validation design adequately tests generalization is itself an open methodological question (Varoquaux, 2018).

This problem has direct clinical relevance: objective pain and analgesia monitoring remains unsolved. Pain is a multidimensional experience (Baliki and Apkarian, 2015; Raja et al., 2020), self-report is unavailable in non-verbal, sedated, or cognitively impaired patients (Herr et al., 2011), and current physiological monitors lack specificity for cortical pain processing (Karunakaran et al., 2021). Whether local anesthesia has succeeded – whether the patient’s cortical pain response has been suppressed – is something clinicians can only judge indirectly, with the economic burden of inadequate pain management estimated at US$560–635 billion annually in the United States alone (Gaskin and Richard, 2012).

Portable functional near-infrared spectroscopy (fNIRS) can measure hemodynamic changes at the bedside (Ferrari and Quaresima, 2012; Hu et al., 2021). Noxious stimuli activate contralateral primary somatosensory cortex (S1) and prefrontal cortex (PFC) in patterns detectable by fNIRS (Aasted et al., 2016; Yucel et al., 2015), including under pharmacological analgesia (Green et al., 2022; Peng et al., 2018). Machine-learning decoding of pain from fNIRS has been explored in primary studies (Fernandez Rojas et al., 2024; Fernandez Rojas et al., 2019; Hu et al., 2019; Zeng et al., 2023) and reviews (Hall et al., 2021; Hu et al., 2021). Our group previously demonstrated real-time pain detection using a neural-network framework with within-session cross-validation (Hu et al., 2019), achieving promising accuracy. However, within-session validation can yield optimistic estimates and does not test subject-independent generalization (Varoquaux, 2018), and strict subject-independent validation has rarely been applied to clinical pain stimuli with pharmacological verification of analgesic state. How large the gap between within-session and cross-patient performance is – and whether per-patient calibration can bridge it – has not been quantified.

Two approaches may address the between-individual scaling problem. First, per-patient calibration: recording a brief baseline from each patient to normalize their brain responses before classification, an approach with precedent in EEG-based pain prediction using spontaneous-activity normalization (Bai et al., 2016) and analogous to per-subject normalization in EEG-based brain-computer interfaces. Second, identifying features that are inherently more stable across individuals: recent EEG evidence suggests that trial-to-trial neural variability encodes interindividual differences in pain discriminability independently of evoked amplitude (Zhang et al., 2025), and deoxyhemoglobin (HbR) may be less affected by extracerebral confounds than oxyhemoglobin (HbO) (Tachtsidis and Scholkmann, 2016). Whether these signals translate to clinical fNIRS pain detection with pharmacological verification has not been tested.

We addressed these questions using fNIRS data acquired during local nerve anesthesia in patients with clinically painful teeth, aiming to (1) characterize the within-patient hemodynamic analgesia signature; (2) test whether per-patient calibration with those features enables permutation-validated analgesic-state detection under strict leave-one-subject-out (LOSO) evaluation; and (3) quantify the inflation from within-session cross-validation, including for end-to-end deep learning on raw fNIRS signals.

## 2. Methods

### 2.1. Participants and analytic sample

The study was approved by the University of Michigan Institutional Review Board, and all participants provided written informed consent. Subjects were recruited at the Endodontics clinic, University of Michigan School of Dentistry. Eligibility required age 18 to 65 years, no ongoing medical or mental health conditions, and a clinical diagnosis of symptomatic apical periodontitis following American Association of Endodontists criteria. Participants were additionally required to have a healthy contralateral tooth. Exclusion criteria included chronic pain disorders, psychotic or severe depressive disorders, disability litigation, neurological disorders, or any severe condition interfering with the study. Participants abstained from analgesics for at least 6 hours before recording. Each participant underwent two fNIRS recording sessions – "Pre" and "Post" local nerve anesthesia. Each session comprised: (1) 5-minute resting-state monitoring; (2) a percussion stimulus block with 8 trials to the contralateral healthy tooth ("NoPain" condition) followed by 8 trials to the affected painful tooth ("Pain" condition), separated by approximately 10-second inter-trial intervals. Local anesthetic was 4% articaine with 1:200,000 epinephrine (Septodont, Saint-Maur-des-Fosses, France) via buccal infiltration (maxillary) or inferior alveolar nerve block (mandibular), preceded by 20% benzocaine topical formula.

Thirty patients were enrolled (17 female, 13 male). Five were excluded: two (subjects 1 and 16) had no fNIRS data acquired, and three (subjects 5, 6, and 8) had all recording sessions fail quality control (fewer than 50% of channels valid). This left 25 subjects with usable data. Of the 30 enrolled, 21 had confirmed successful local nerve anesthesia (block-success), 6 had persistent pain response indicating incomplete block (block-failure), and anesthesia outcome was unavailable for the remaining 3; of the block-failure subjects, 4 had usable data and were retained for held-out validation. The effective sample varied across analyses: 13 block-success subjects with complete Pre/Post Pain data entered the within-patient interaction and feature-based calibrated classification analyses, 23 entered the cross-patient pain-detection analysis (22 with both Pain and NoPain epochs in the variability comparison), 17 block-success subjects with valid connectivity data entered the regional connectivity analysis, and 13 block-success and 4 block-failure subjects with Pre-session data entered the block-outcome prediction analysis (n = 17). After quality exclusion, 626 percussion events were retained (NoPain: n = 372; Pain: n = 254).

### 2.2. fNIRS data acquisition and signal processing

Data were acquired using a continuous-wave system (NIRSport2, NIRx Medical Technologies, Glen Head, NY) with 8 sources and 8 detectors, yielding 18 source-detector pairs covering PFC (4 pairs) and bilateral S1 (7 pairs per hemisphere). Each pair provided separate HbO and HbR signals, giving 36 channels per event (18 x 2).

Signal processing proceeded in three stages. Stage 1: channels with fewer than 5% missing samples were reconstructed by cubic spline interpolation; those with 5% or more were excluded. Channels were automatically pruned on the basis of cardiac pulsation (0.5–2.0 Hz). Signal quality was quantified using the Scalp Coupling Index (SCI) (Pollonini et al., 2016) and a custom Cardiac Peak Signal-to-Noise Ratio (CpSNR), defined as the ratio of peak cardiac spectral power to the aperiodic background estimated via the FOOOF approach (Donoghue et al., 2020). Channels were retained only if SCI > 0.5 and CpSNR > 1.2. Stage 2: optical density conversion, motion correction via Temporal Derivative Distribution Repair with principal component analysis followed by wavelet-based filtering (3-sigma threshold), and modified Beer-Lambert law conversion to HbO and HbR concentration changes (NIRS Brain AnalyzIR Toolbox). Sampling rates were 7.8125 Hz (subjects 1–22) and 10.1725 Hz (subjects 23–30); all data were resampled to 8 Hz. Stage 3: recordings with fewer than 50% valid channels were dropped (10 of 56 sessions). After remapping channels to contralateral/ipsilateral designations based on affected tooth side, all 36 channel slots were carried into feature extraction (11 features × 36 channels = 396 features), with absent channels zero-filled. Of these, 24 channels were well-populated (data in >50% of epochs: 2 contralateral S1, 14 ipsilateral S1, 8 PFC), 6 contralateral S1 channels were sparsely populated (data in 12–14% of epochs), and 6 were globally absent (valid in <50% of recordings). End-to-end deep learning operated on the 30 channels with any valid data (72 timepoints × 30 channels), excluding the 6 globally absent channels.

### 2.3. Feature extraction

For each percussion event, a hemodynamic response window spanning 1 s pre-stimulus to 8 s post-stimulus was extracted and baseline-corrected to the mean of the final 0.75 s of the pre-stimulus period. Windows with fill ratio below 0.75 were discarded.

Eleven feature types were computed per channel per event: GLM response amplitude (beta), model fit (R-squared), peak latency, onset latency, full-width at half-maximum (FWHM), baseline-normalized amplitude (z-scored mean), mean first derivative, late slope, baseline mean, wavelet noise energy ratio, and trough amplitude (z-scored trough) – 396 features total. The GLM used a canonical double-gamma hemodynamic response function (Glover, 1999). Six epoch-level functional connectivity summary statistics (mean and maximum pairwise correlation, partial correlation) were also computed. For the variability analysis, trial-to-trial standard deviation of each feature was calculated within each (subject, session, stimulus type) group, yielding a matched set of 396 variability features.

### 2.4. Classification and statistical analysis

Analyses were organized into primary and additional tiers to manage multiple-testing concerns. The three primary analyses were: (1) within-patient analgesia signature, in which the interaction contrast Delta = (Pain_Pre – NoPain_Pre) – (Pain_Post – NoPain_Post) was computed per subject for each of 11 feature types and tested with one-sample t-tests; (2) calibrated analgesic-state classification under LOSO, using features identified by the within-patient analysis, with 500-permutation within-subject label-shuffle testing, a negative control on healthy-tooth epochs, and available block-failure data as a secondary check; and (3) validation inflation, comparing within-session stratified 5-fold CV (repeated 20 times) with strict LOSO on the same data, features, and classifiers. The aggregate within-patient effect was additionally tested with a difference-in-differences (DiD) model (Angrist and Pischke, 2009): a subject-demeaned ordinary least squares regression (feature ∼ Pain + Post + Pain x Post) in which the Pain x Post interaction term estimates the analgesia-specific change after removing main effects of session and stimulus type, with the healthy tooth serving as the within-subject control for session-level drift. Cluster-bootstrap confidence intervals (resampling subjects, 5000 iterations) were used to account for within-subject correlation.

Per-patient calibration consisted of z-scoring each patient’s features to their own Pre-session NoPain epoch statistics (mean and SD), removing between-subject scaling while preserving the Pain-versus-NoPain and Pre-versus-Post contrasts. The calibrated classifier outputs a continuous score for each epoch: values near 1 indicate a Post (pain-abolished) cortical pattern, values near 0 indicate a Pre (pain-present) pattern; the threshold for classification is 0.5. The feature-based classification framework encompassed logistic regression (C = 0.1), linear discriminant analysis (LDA), radial basis function support vector machine (RBF-SVM), and random forest (RF), with automated principal component analysis (PCA) when dimensionality exceeded the training sample size.

As a state-of-the-art comparison, three end-to-end deep learning architectures commonly used in fNIRS (Fernandez Rojas et al., 2024) and electroencephalography (EEG) brain-computer interface studies were applied directly to raw baseline-corrected fNIRS time series (72 timepoints x 30 channels at 8 Hz) under the same LOSO and within-session CV framework: (1) a 1D convolutional neural network (CNN1D; three Conv1d layers with 64, 128, and 64 filters, kernel sizes 7, 5, and 3, batch normalization, max-pooling, and adaptive global average pooling, followed by a fully connected head with 32 hidden units and 50% dropout); (2) a bidirectional long short-term memory network (BiLSTM; 2-layer BiLSTM with 64 hidden units per direction, 30% recurrent dropout, followed by a 32-unit fully connected head with 50% dropout); and (3) a CNN-LSTM hybrid (two Conv1d layers with 64 filters each, kernel sizes 5 and 3, feeding into a 1-layer BiLSTM with 32 hidden units, followed by the same fully connected head). All models were trained with the Adam optimizer (learning rate 1e-3, weight decay 1e-4), class-weighted binary cross-entropy loss, and early stopping on a stratified 15% validation split. For the deep learning models, per-patient calibration consisted of subtracting each patient’s mean Pre-NoPain HRF waveform and dividing by its standard deviation – the signal-level analogue of the feature-based z-score calibration. Data augmentation (Gaussian noise at 5% of channel standard deviation, temporal jitter of +/-2 samples, channel-wise scaling drawn from uniform [0.9, 1.1]; 4 augmented copies) was applied only inside training folds, and dead channels were zero-filled with explicit channel masks to prevent the model from exploiting missingness patterns as subject identifiers. Hyperparameters were fixed to standard values from the literature without tuning. Permutation testing for deep learning models used 50 within-subject label shuffles (vs 500 for feature-based models) due to computational cost.

Additional analyses included uncalibrated classification (to assess the necessity of calibration), cross-patient pain detection (Pre session, Pain vs NoPain), chromophore decomposition of the within-patient interaction and calibrated classification, variability versus amplitude comparison (Bayesian Beta-binomial and exact McNemar test), block-outcome prediction, and regional connectivity.

## 3. Results

### 3.1. Dataset

After quality exclusion (10 of 56 sessions dropped for insufficient valid channels; subjects 5, 6, and 8 lost entirely; additional epochs discarded for window fill ratio below 0.75), 626 valid percussion windows across 46 sessions from 25 subjects constituted the analytic dataset. Thirteen block-success subjects had complete data in all four cells of the Pre/Post x Pain/NoPain design (156 pain epochs: 77 Pre, 79 Post; 213 NoPain epochs: 108 Pre, 105 Post). Between-subject heterogeneity in pain-evoked amplitude was substantial (Figure 1), with some subjects showing larger responses to the healthy tooth than to the painful one, underscoring the need for individual calibration.

**Figure 1.**
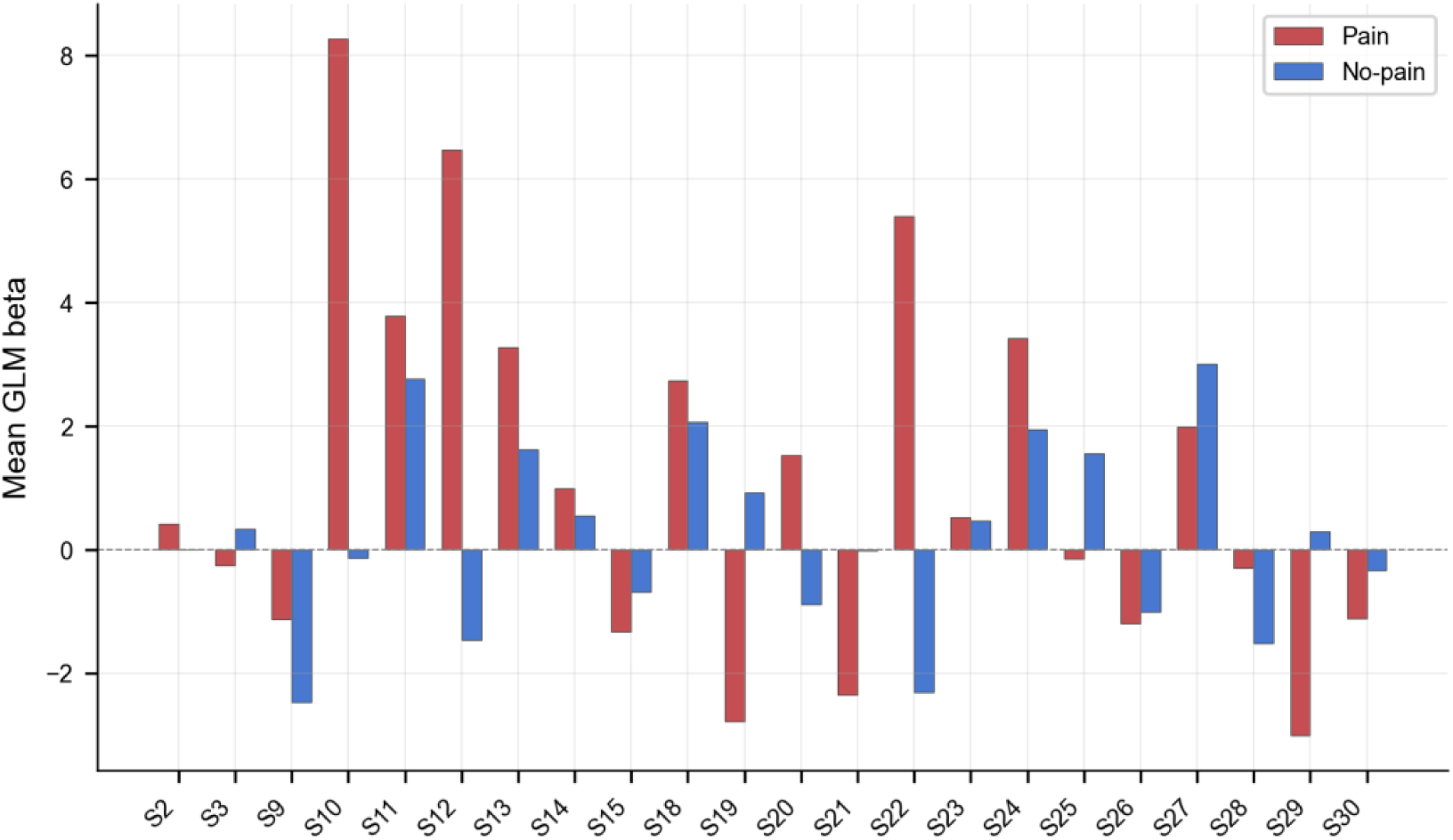
Per-subject mean GLM response amplitude for pain (red) and no-pain (blue) in the Pre session. Most subjects show larger pain responses, but the magnitude and direction vary substantially – some show the reverse pattern – motivating per-patient calibration.

### 3.2. Within-patient hemodynamic analgesia signature

The within-patient analgesia interaction (n = 13 block-success patients) revealed that the temporal dynamics of the hemodynamic response were more consistently affected by anesthesia than its peak magnitude (Figure 2, Table 1). Four features reached significance: late slope (d = 0.79, p = 0.015), baseline-normalized amplitude (d = 0.73, p = 0.022), trough amplitude (d = 0.70, p = 0.028), and mean first derivative (d = 0.63, p = 0.044). Raw GLM amplitude showed a medium effect that did not reach significance (d = 0.56, p = 0.065). Three features were selected for the calibrated classifier – late slope, baseline-normalized amplitude, and mean first derivative – because they capture complementary temporal information (rate of HRF rise, sustained profile, and normalized magnitude). Trough amplitude, despite its larger effect size, was excluded because it was highly correlated with baseline-normalized amplitude (r = 0.56), whereas mean first derivative was near-orthogonal to both amplitude measures (r < 0.1). The aggregate epoch-level model (below) confirmed the overall effect.

**Figure 2.**
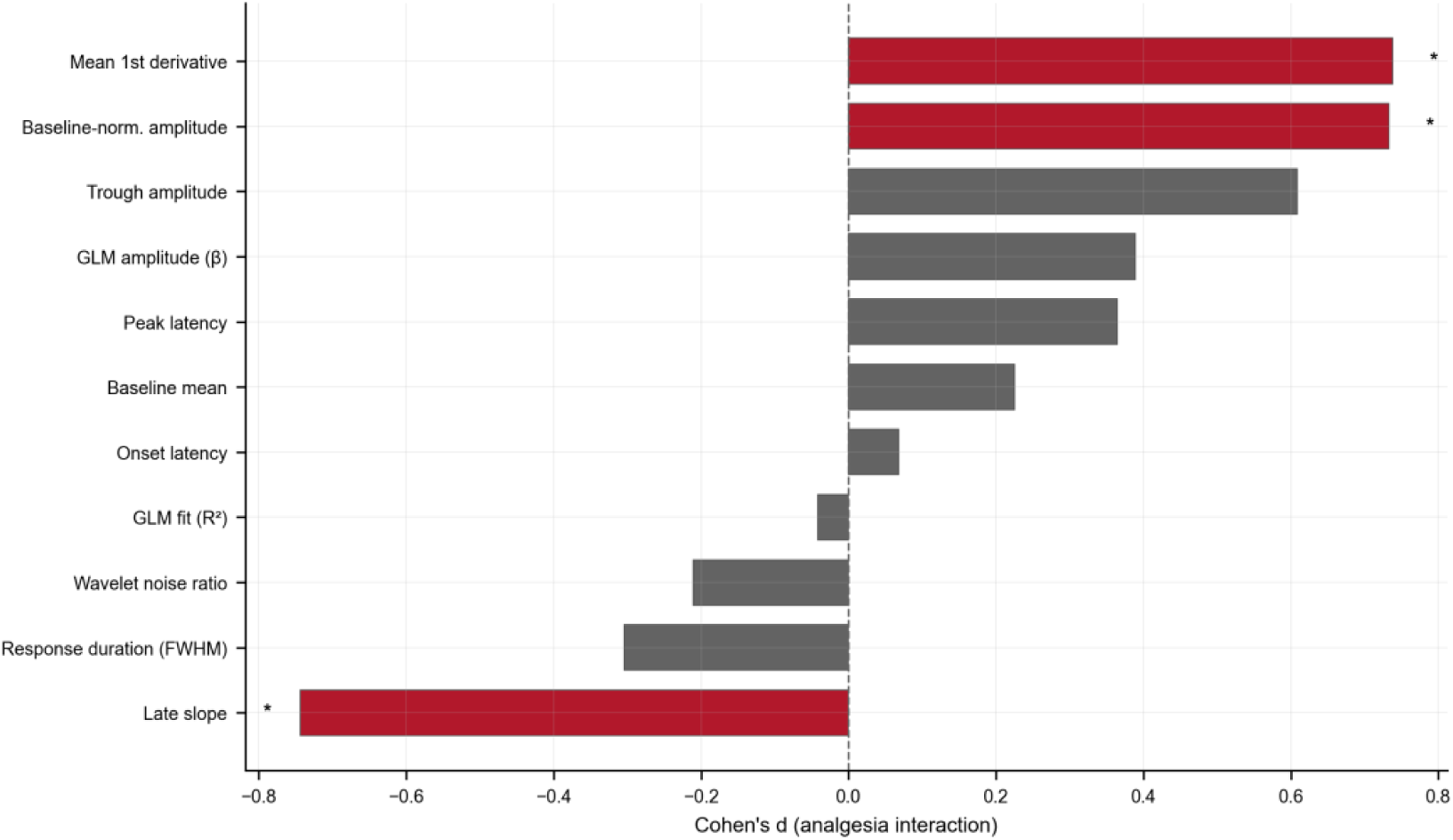
Within-patient analgesia interaction effects (n = 13). Cohen’s d for each of 11 HRF feature types. Four features reach significance (starred); three were selected for the calibrated classifier (late slope, baseline-normalized amplitude, mean first derivative) based on complementary temporal information (see text). Raw GLM amplitude shows a medium non-significant effect (d = 0.56, grey).

**Table 1.** Within-patient analgesia interaction by HRF feature type (n = 13; one-sample t-test on Delta = [Pain_Pre - NoPain_Pre] - [Pain_Post - NoPain_Post]).

| Feature | Cohen d | p |
| --- | --- | --- |
| Late slope | -0.79 | 0.015 |
| Baseline-norm. amplitude | +0.73 | 0.022 |
| Trough amplitude | +0.70 | 0.028 |
| Mean 1st derivative | +0.63 | 0.044 |
| GLM amplitude (beta) | +0.56 | 0.065 |
| FWHM | -0.43 | 0.143 |
| Wavelet noise ratio | -0.32 | 0.278 |
| Peak latency | +0.30 | 0.293 |
| GLM fit (R-squared) | -0.21 | 0.465 |
| Baseline mean | +0.17 | 0.554 |
| Onset latency | +0.03 | 0.905 |

The DiD model (see Methods) fitted on the mean of the top-3 features across 410 epochs from all 15 block-success subjects with both Pre and Post sessions (a larger set than the 13 entering the calibrated classifier, which additionally required ≥2 pain epochs per session) confirmed the aggregate effect: the Pain x Post interaction was significant (coefficient = −1.11; cluster-bootstrap 95% CI [−2.07, −0.22], p = 0.011), indicating that the pain-evoked hemodynamic contrast was reduced after anesthesia, net of any session effect on the healthy tooth. This pattern suggests that local anesthesia suppresses the temporal unfolding of the cortical pain response – its rate of rise and sustained profile – more reliably than it reduces peak amplitude.

### 3.3. Calibrated analgesic-state classification

The calibrated classifier was designed to address a clinical question: after local anesthesia, does the patient’s brain still show a pain response? Each patient is first calibrated with a brief Pre-anesthesia percussion block, during which both the painful and healthy teeth are tapped. The classifier learns that patient’s cortical Pain-versus-NoPain contrast and then evaluates Post epochs to determine whether the pain pattern persists or has been suppressed. Because the classifier is trained on Pre-versus-Post session labels rather than subjective pain reports, a negative control is important: the same pipeline applied to healthy-tooth epochs should show no Pre-versus-Post difference.

Per-patient calibration markedly improved LOSO classification (Figure 3, Table 2). With the top-3 temporal features across both chromophores (108 features), nonlinear classifiers reached significance: RF AUC = 0.705 (SD = 0.271, p = 0.002) and RBF-SVM AUC = 0.679 (SD = 0.148, p = 0.002), while linear models (LDA, logreg) did not exceed the permutation null. Restricting to HbO channels only (54 features) further improved the best model: RF reached AUC = 0.760 (SD = 0.207), balanced accuracy = 0.722, p = 0.002; RBF-SVM reached AUC = 0.701 (SD = 0.176, p = 0.002). The improvement from chromophore-specific feature selection (HbO-only RF: +0.055 AUC over both-chromophore RF) suggests that HbR channels introduce noise in the classification task despite carrying a within-patient analgesia signal (see Section 3.5).

**Figure 3.**
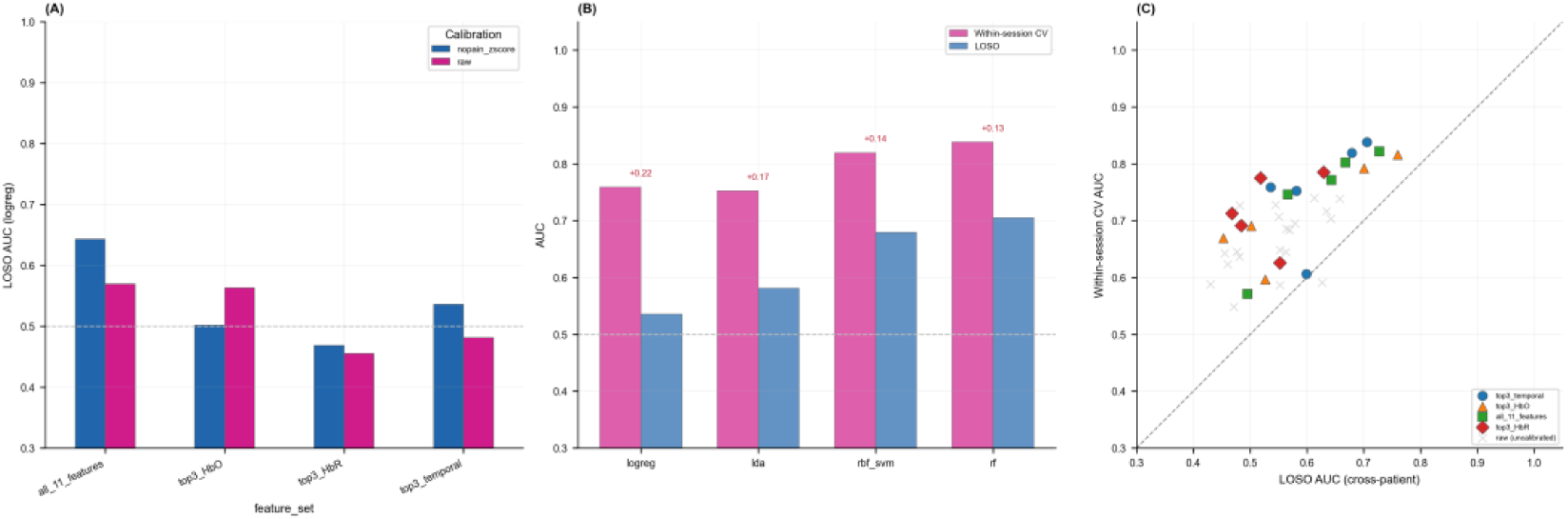
Calibrated analgesic-state classification and validation inflation. (A) Effect of calibration: LOSO AUC for calibrated (blue) versus raw (pink) features across feature sets. (B) Validation inflation for calibrated top-3 temporal features: within-session CV (pink) versus LOSO (blue) AUC, with inflation annotated. (C) Inflation scatter across all configurations (above diagonal = inflated).

**Table 2.** Calibrated LOSO analgesic-state classification (Pre vs Post pain epochs, n = 13 subjects). Feature-based models: NoPain z-scored top-3 temporal features, 500-permutation test. DL models: raw time series with HRF-subtraction calibration.

| Input | Model | AUC mean +/- SD | BAcc | Perm p |
| --- | --- | --- | --- | --- |
| Top-3 temporal, both (108) | RF | 0.705 +/- 0.271 | 0.703 | 0.002 |
| Top-3 temporal, both (108) | RBF-SVM | 0.679 +/- 0.148 | 0.664 | 0.002 |
| Top-3 temporal, both (108) | LDA | 0.581 +/- 0.247 | 0.580 | 0.074 |
| Top-3 temporal, both (108) | Logreg | 0.536 +/- 0.293 | 0.537 | 0.297 |
| Top-3 temporal, HbO only (54) | RF | 0.760 +/- 0.207 | 0.722 | 0.002 |
| Top-3 temporal, HbO only (54) | RBF-SVM | 0.701 +/- 0.176 | 0.679 | 0.002 |
| Raw signal (72x30) | CNN-LSTM | 0.719 +/- 0.171 | 0.620 | 0.020 |
| Raw signal (72x30) | BiLSTM | 0.625 +/- 0.189 | 0.604 | 0.020 |
| Raw signal (72x30) | CNN1D | 0.616 +/- 0.243 | 0.563 | 0.020 |

End-to-end deep learning on raw time series, with HRF-subtraction calibration, also achieved above-chance LOSO performance. Notably, CNN-LSTM (AUC = 0.719, SD = 0.171) matched or exceeded the feature-based classifiers using the default top-3 temporal features across both chromophores (RF AUC = 0.705), though it fell below HbO-specific feature selection (RF AUC = 0.760). This indicates that end-to-end learning on raw signals can partially compensate for suboptimal feature selection, while domain knowledge applied to chromophore selection still provides the best overall performance. The observation that linear models (logreg, LDA) failed to reach significance while nonlinear models (RF, RBF-SVM) and deep learning succeeded suggests that the analgesic-state boundary in feature space is nonlinear.

As a negative control, applying the same calibrated pipeline to healthy-tooth (NoPain) epochs produced near-chance LOSO performance across all classifiers, confirming that the classifier detects pain-specific analgesia rather than session-level drift.

For context, the same calibrated HbO-RF yielded within-session AUC = 0.816, comparable to the best published fNIRS pain-decoding results (Fernandez Rojas et al., 2024; Fernandez Rojas et al., 2019; Hu et al., 2019). The LOSO result (0.760) is the stricter cross-patient benchmark.

To address the circularity of selecting features on the same subjects used for classification, we repeated the analysis with per-fold feature selection: in each LOSO fold, the within-patient interaction was recomputed on training subjects only and the top-3 features by effect size were selected. Nested LDA achieved AUC = 0.566 (SD = 0.248), with the same three features selected in the majority of folds. Repeating the analysis with only the 24 well-populated channels (72 features instead of 108), excluding the 12 zero-filled contralateral S1 channels, yielded similar results (LDA AUC = 0.428, logreg AUC = 0.508), confirming that the findings are not driven by the zero-filled channels.

### 3.4. Validation inflation

A head-to-head comparison of within-session CV and LOSO on the analgesic-state task quantified the inflation (Figure 3, Table 3). For calibrated temporal features, within-session CV inflated AUC by +0.06 to +0.14 across the significant feature-based classifiers. HbO-specific feature selection reduced inflation (HbO-RF: +0.057) compared with both-chromophore features (RF: +0.133), consistent with HbO features being more stable across patients. Without calibration, inflation was larger: raw features yielded within-session AUC = 0.727 but LOSO AUC = 0.482 – an inflation of +0.25.

**Table 3.**
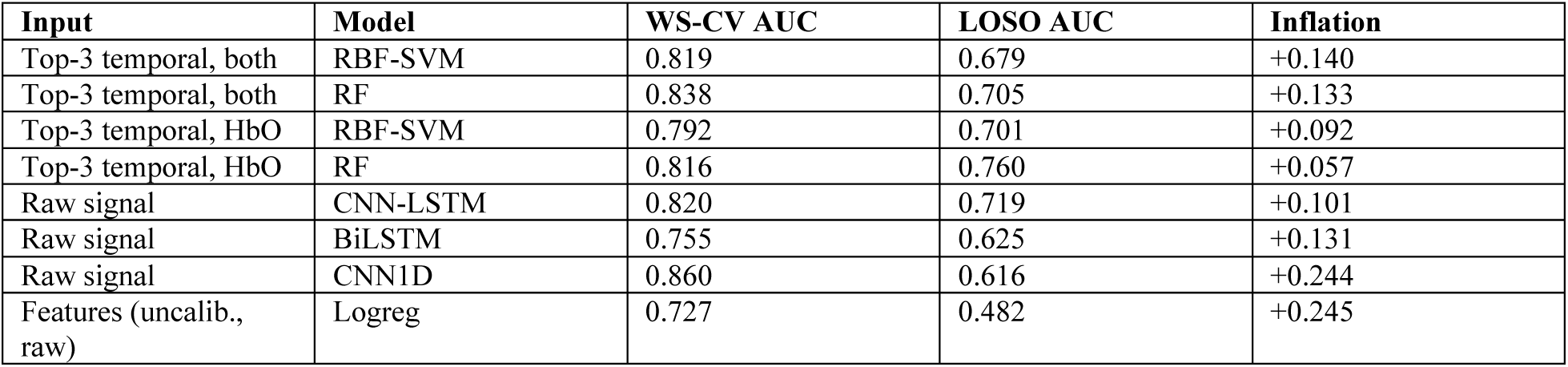
Validation inflation: within-session CV versus LOSO (n = 13). Feature-based models: calibrated top-3 temporal features. DL models: calibrated raw signal.

The same pattern held for deep learning: CNN1D showed +0.24 inflation (within-session 0.860 vs LOSO 0.616); BiLSTM +0.13 (0.755 vs 0.625); CNN-LSTM +0.10 (0.820 vs 0.719). Across all configurations, the mean inflation was +0.13 AUC.

Within-session CV conflates within-person and between-person variance, producing estimates that do not transfer across patients. This inflation is not specific to the feature-extraction step; it occurs with end-to-end deep learning on raw time series as well. Per-patient calibration and targeted feature selection jointly reduce the gap, though they do not eliminate it entirely.

### 3.5. Additional analyses

We report several additional analyses that provide converging evidence and may inform future studies with larger samples.

Without per-patient calibration, classification was at chance under LOSO (logreg AUC = 0.48, LDA AUC = 0.46). Calibration lifted performance to 0.68–0.76, confirming that between-subject scaling must be addressed for fNIRS pain decoding to generalize. Classifying Pain versus NoPain from Pre epochs alone (305 epochs, 23 subjects) – a harder task because it lacks per-patient calibration – yielded modest LOSO performance (Table 4): RF AUC = 0.591 (SD = 0.193) was the strongest feature-based result, and CNN1D AUC = 0.591 (SD = 0.185) was comparable among DL models.

**Table 4.**
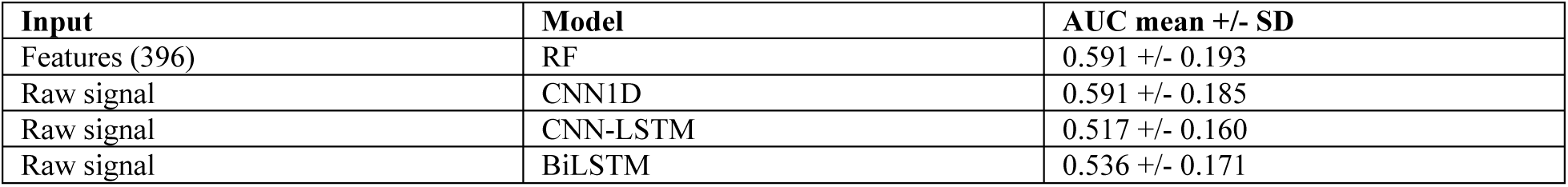
Cross-patient pain detection (Pre session; LOSO; 305 epochs; 23 subjects).

HbO carried the within-patient analgesia signal (d = 1.04, p = 0.003) and dominated the calibrated classifier (HbO-only RF AUC = 0.76), while HbR was not significant overall (d = 0.13) but showed a pronounced PFC-specific effect (d = 0.66, p = 0.035; Figure 4, Table 5). These results suggest complementary roles for HbO and HbR, and caution against relying on HbO alone, especially given the literature’s strong emphasis on HbO changes (Hall et al., 2021).

**Figure 4.**
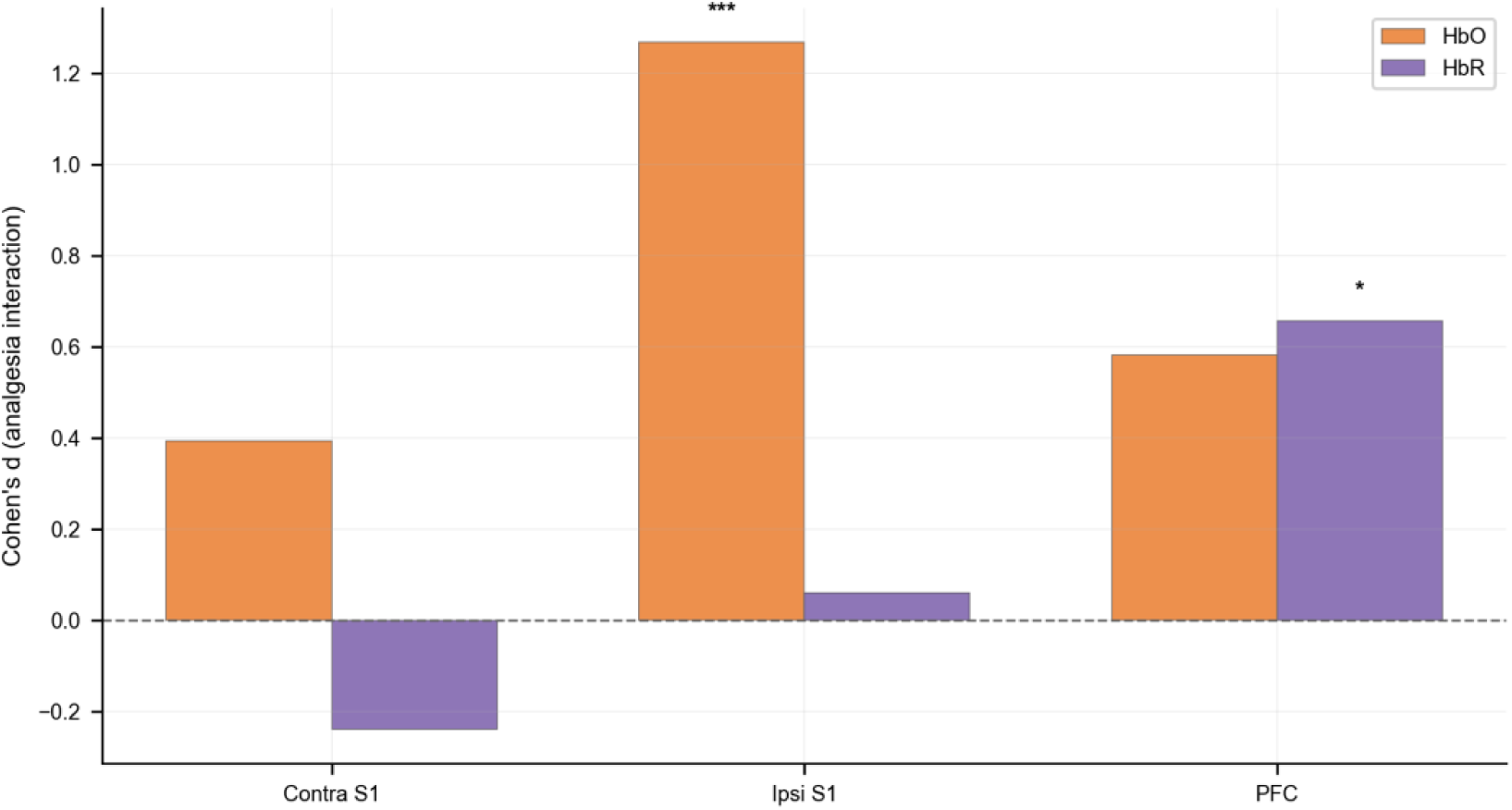
Chromophore x region analgesia interaction (n = 13). Cohen’s d for HbO (orange) and HbR (purple) across contralateral S1, ipsilateral S1, and PFC. HbO shows broad analgesia sensitivity (d = 1.04, p = 0.003); HbR shows a PFC-specific effect (d = 0.66, p = 0.035).

**Table 5.** Chromophore x region analgesia interaction (n = 13; one-sample t-test).

| Chromophore | Region | Cohen d | p |
| --- | --- | --- | --- |
| HbO | Overall | +1.04 | 0.003 |
| HbO | Contra S1 | +0.39 | 0.180 |
| HbO | Ipsi S1 | +1.27 | 0.001 |
| HbO | PFC | +0.58 | 0.058 |
| HbR | Overall | +0.13 | 0.639 |
| HbR | Contra S1 | -0.24 | 0.408 |
| HbR | Ipsi S1 | +0.06 | 0.829 |
| HbR | PFC | +0.66 | 0.035 |

Trial-to-trial variability features classified pain above chance at the subject level (16/22 subjects with both Pain and NoPain Pre data correctly classified, binomial p = 0.026), while amplitude did not (13/22, p = 0.26; Table 6). The direct comparison was inconclusive (Bayesian posterior 82%, 95% credible interval [−0.14, +0.38], McNemar p = 0.25 on 9 discordant subjects), but the trial-degradation curve was informative: variability AUC rose monotonically from chance at k = 2 trials to 0.83 at k = 8, while amplitude remained flat near 0.60 (Figure 5). This accumulation pattern – more trials yielding better discrimination – is the expected signature of a consistency-based signal and parallels recent EEG findings on neural variability and pain discriminability (Zhang et al., 2025).

**Figure 5.**
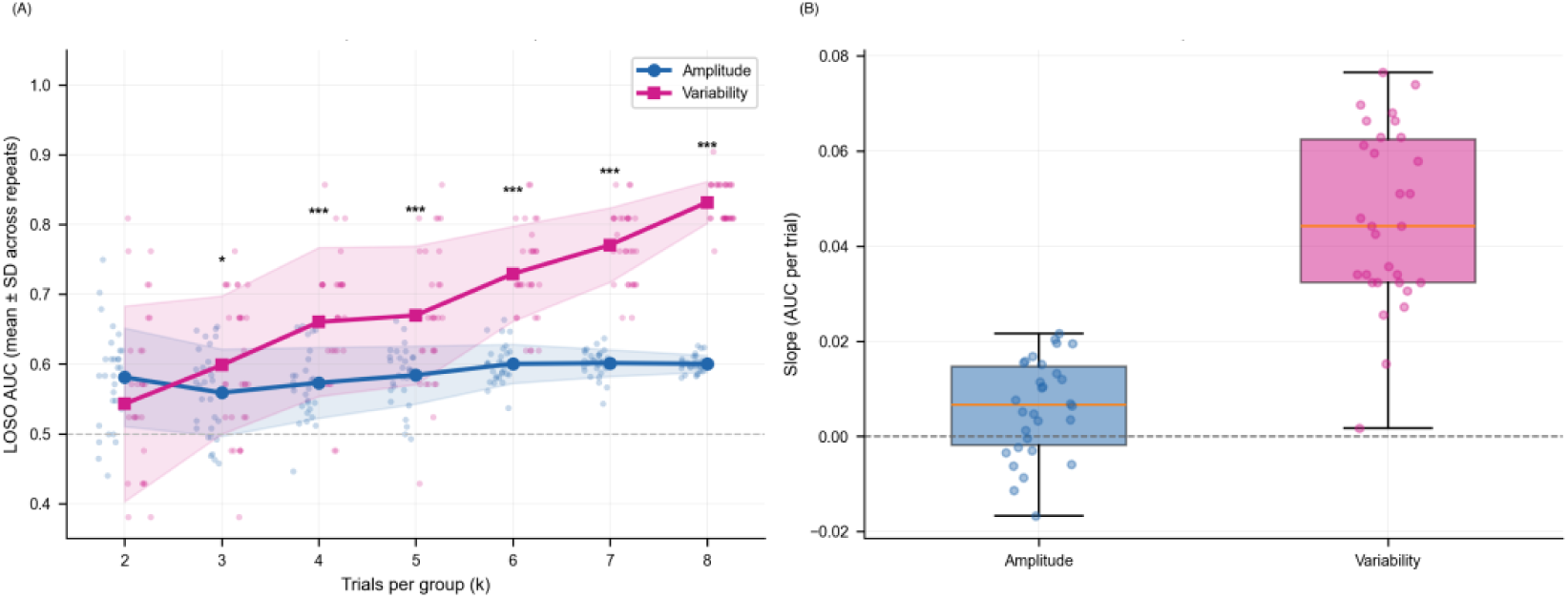
Trial-degradation curve with statistical tests (Pre session; LOSO; 30 sub-sampling repeats per k). (A) Degradation curve: variability AUC (pink) rises monotonically from chance at k = 2 to 0.83 at k = 8 trials; amplitude AUC (blue) remains flat near 0.60. Stars indicate paired t-tests (variability > amplitude) at each k. Error bands: +/- 1 SD across repeats. (B) Per-repeat AUC-vs-k slopes: variability slopes are consistently positive while amplitude slopes cluster near zero (paired t, p < 0.0001, d = 2.0), confirming that variability accumulates diagnostic information with increasing trial count whereas amplitude does not.

**Table 6.**
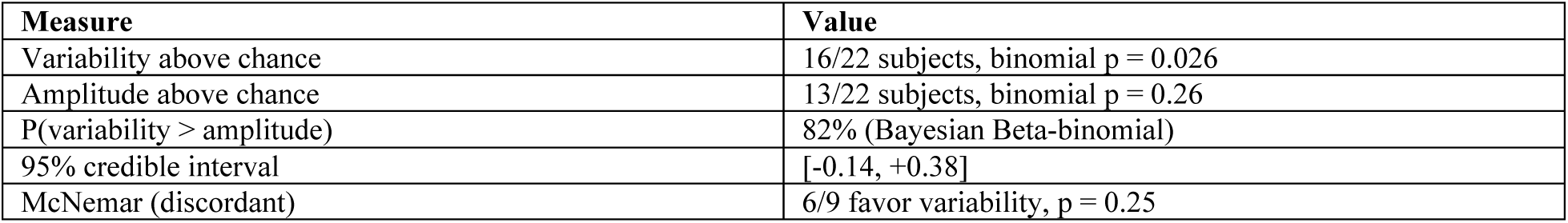
Cross-patient variability versus amplitude (Pre session; LOSO; 396 features each; 22 subjects with both pain and no-pain Pre-session data).

| Measure | Value |
| --- | --- |
| Variability above chance | 16/22 subjects, binomial $p = 0.026$ |
| Amplitude above chance | 13/22 subjects, binomial $p = 0.26$ |
| $P(\text{variability} > \text{amplitude})$ | 82% (Bayesian Beta-binomial) |
| 95% credible interval | [-0.14, +0.38] |
| McNemar (discordant) | 6/9 favor variability, $p = 0.25$ |

Regional connectivity in the 17 block-success subjects with valid connectivity data showed contralateral S1–ipsilateral S1 coupling declining after anesthesia (d = 0.69, p = 0.011) as did contralateral S1–PFC coupling (d = 0.58, p = 0.035), while ipsilateral S1–PFC coupling showed a trend (d = 0.50, p = 0.057; Figure 6, Table 7).

**Figure 6.**
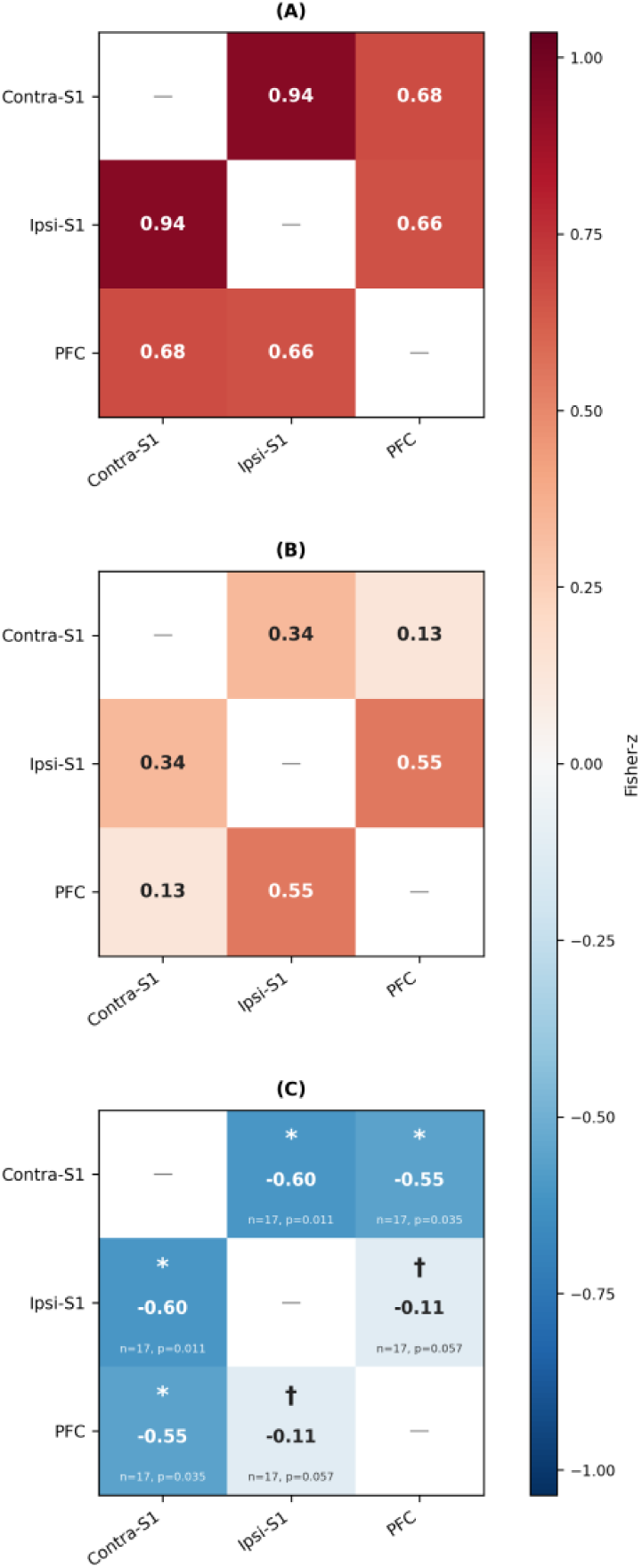
Regional fNIRS connectivity for pain events (n = 17 block-success subjects, uncorrected). (A) Pre-anesthesia Fisher-z correlation matrix. (B) Post-anesthesia Fisher-z correlation matrix. (C) Difference (Post – Pre) in Fisher-z units. Paired Wilcoxon tests: contralateral S1–ipsilateral S1 d = 0.69, p = 0.011; contralateral S1–PFC d = 0.58, p = 0.035; ipsilateral S1–PFC d = 0.50, p = 0.057. Cell values in (C) are group-mean Fisher-z differences; Cohen’s d and p values are from paired within-subject tests.

**Table 7.**
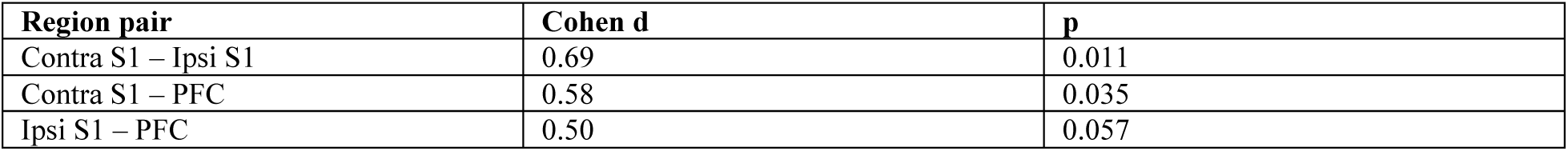
Regional connectivity Pre vs Post (pain events; n = 17 block-success subjects with valid connectivity data; paired Wilcoxon, uncorrected).

Cortical surface projections show selective contralateral S1 suppression (Figure 7), and grand-average HRF time courses illustrate the amplitude reduction (Figure 8). A separate question is whether block outcome can be predicted from Pre data alone, before anesthesia is administered. Pre-procedure contralateral S1 variability was higher in block-success than block-failure subjects (d = 1.15, p = 0.032; 13 block-success vs 4 block-failure, n = 17 total; Figure 9), while PFC variability and S1 amplitude were not predictive. The effect size is large, but post-hoc power was only 0.47 – a lead that requires a larger sample.

**Figure 7.**
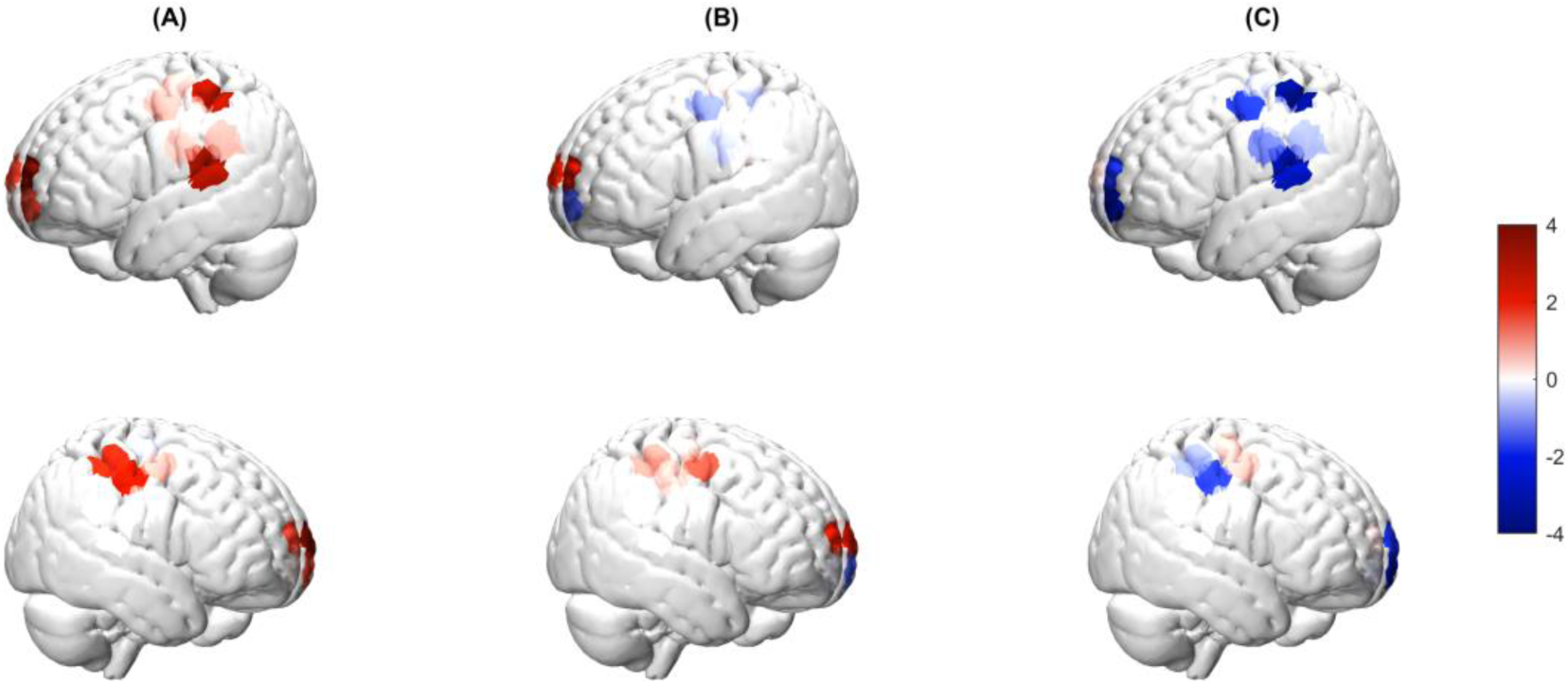
Cortical surface projection of group-mean GLM-beta (HbO). Top row: lateral view; bottom row: dorsal view. (A) Pre-anesthesia: contralateral S1 activation. (B) Post-anesthesia: reduced activation. (C) Difference (Post – Pre): selective contralateral S1 and PFC suppression (blue).

**Figure 8.**
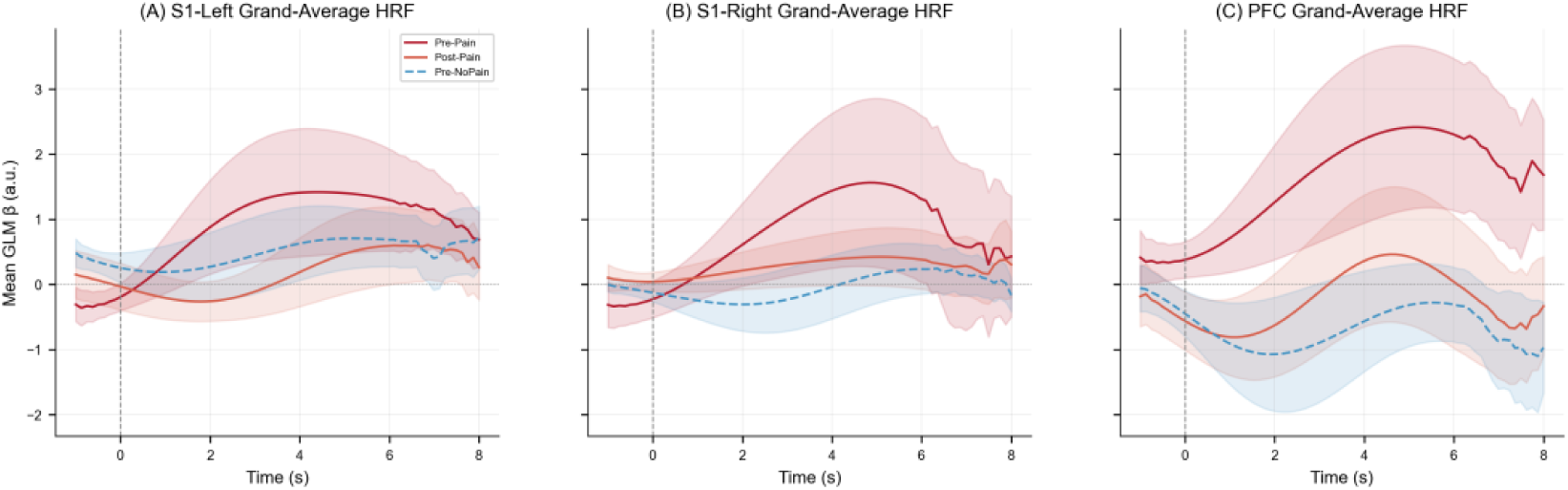
Grand-average HRF time courses for (A) S1-left, (B) S1-right, and (C) PFC. Traces show Pre Pain (solid red), Post Pain (dark red), Pre NoPain (solid blue), and Post NoPain (dashed blue); shading indicates +/- 1 SEM. Vertical dashed line marks stimulus onset.

**Figure 9.**
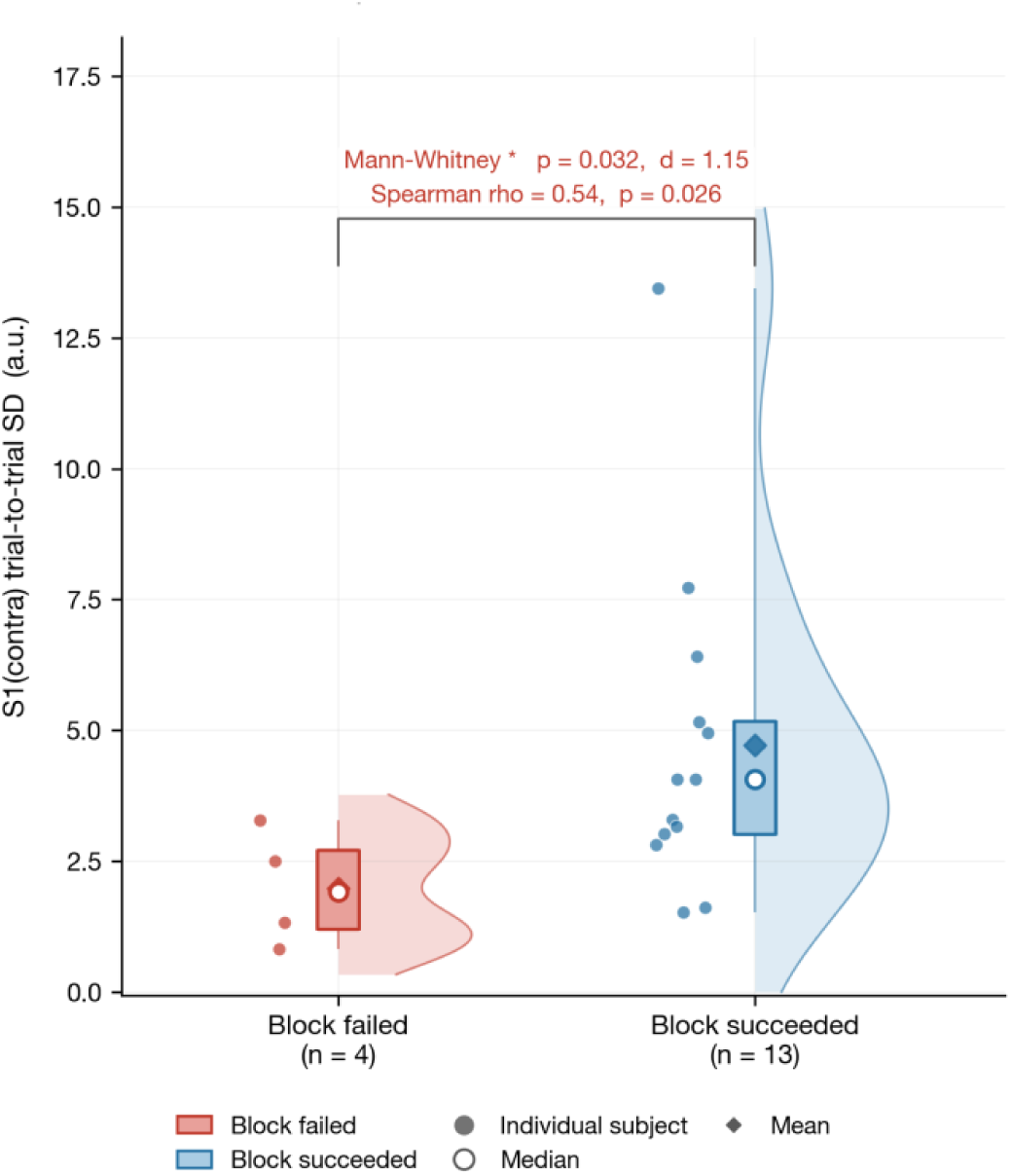
Pre-procedure contralateral S1 trial-to-trial variability by block outcome (n = 17 total). Block-success (blue, n = 13) > block-failure (red, n = 4; Mann-Whitney p = 0.032, d = 1.15; Spearman rho = 0.54, p = 0.026). Individual subjects shown as dots; diamonds indicate group means; circles indicate medians.

## 4. Discussion

### 4.1. Main findings

Per-patient calibration with three temporal HRF features yielded subject-independent analgesic-state detection under strict LOSO evaluation. The best performance was achieved by combining calibration with chromophore-specific feature selection: HbO-only RF reached LOSO AUC = 0.760 (p = 0.002), followed by HbO-only RBF-SVM at 0.701 (p = 0.002). Without calibration, all classifiers were at chance – consistent with the between-person scaling problem documented for fMRI pain signatures (Han et al., 2022) and extending it to portable fNIRS.

The temporal analgesia signature – late slope, first derivative, and baseline-normalized amplitude – was more consistent across patients (d = 0.63–0.79) than raw GLM amplitude (d = 0.56), and the aggregate effect was confirmed by a significant difference-in-differences model (p = 0.011). This suggests that anesthesia does not simply reduce the magnitude of the cortical pain response but reshapes its temporal profile, possibly reflecting the disruption of sustained nociceptive processing rather than a uniform gain reduction. The calibrated HbO-RF (LOSO AUC = 0.76) approaches the best published fNIRS pain-decoding accuracies (Fernandez Rojas et al., 2024; Fernandez Rojas et al., 2019; Hu et al., 2019), though those results relied on within-session cross-validation – the very design our inflation analysis shows can overestimate performance by up to 0.24 AUC. The present result is, to our knowledge, the first to approach comparable accuracy under strict cross-patient (LOSO) evaluation.

Within-session CV inflated performance across all model types, with mean inflation of +0.13 AUC. This inflation extended to end-to-end deep learning on raw fNIRS time series (up to +0.24 for CNN1D). Under strict LOSO evaluation, CNN-LSTM on raw signals (AUC = 0.719) matched or exceeded the default feature-based classifiers using both chromophores (top-3 temporal RF AUC = 0.705), while only chromophore-specific feature selection (HbO-only RF AUC = 0.760) outperformed all DL architectures. This suggests that domain knowledge remains valuable primarily through informed feature selection – specifically, restricting to HbO channels – rather than through feature engineering per se. Linear classifiers (logreg, LDA) did not reach significance under LOSO, while nonlinear models (RF, RBF-SVM) and deep learning did, indicating that the analgesic-state boundary is nonlinear in feature space. This pattern parallels the between-person gap documented for fMRI pain signatures (NPS between-person d = 0.49 vs within-person d = 1.45; Han et al., 2022) and aligns with recent recommendations that neuroimaging pain-decoding studies carefully consider how validation design influences reported performance (Kohoutova et al., 2025). The principle extends beyond fNIRS: any neuroimaging classifier evaluated only within-session risks conflating subject identity with the target signal, a concern shared with EEG-based brain-computer interfaces where per-subject calibration is standard practice. Future fNIRS pain studies should report LOSO performance alongside within-session CV; high within-session accuracy alone does not establish subject-independent validity.

### 4.2. Secondary observations

The chromophore decomposition suggests complementary roles for HbO and HbR: HbO dominated the within-patient analgesia signal (d = 1.04, p = 0.003), while HbR showed a region-specific PFC effect (d = 0.66, p = 0.035) – consistent with prior methodological arguments that HbR may be less affected by extracerebral confounds (Tachtsidis and Scholkmann, 2016). These results caution against relying on HbO alone, especially given the literature’s strong emphasis on HbO changes (Hall et al., 2021).

Trial-to-trial response variability was above chance for cross-patient pain detection while amplitude was not, though the direct comparison was inconclusive. Regional S1 connectivity changes and pre-procedure variability as a predictor of block outcome showed suggestive effects at moderate-to-large effect sizes. These analyses are individually underpowered but together are consistent with the idea that the cortical response to pain is a variable process, and that variability itself may carry diagnostic information. These leads warrant testing in larger, purpose-designed studies.

### 4.3. Limitations and conclusion

The primary limitation is sample size (n = 13 for the within-patient analysis, n = 22-23 for cross-patient analyses). The main findings, however, are supported by convergent evidence: permutation-validated classification across multiple classifiers and architectures, a significant epoch-level DiD interaction, nested feature selection, a pain-specific negative control, and robustness to channel coverage. The within-patient HRF effects, while individually significant, were not corrected for multiple comparisons; however, the same features produced significant permutation-validated classification and a significant aggregate DiD interaction, providing converging evidence that the effects are genuine. Twelve of 36 channels (6 globally absent, 6 sparsely populated) were zero-filled, all in contralateral S1, limiting contralateral interpretation. The multistage design carries residual multiple-comparison risk despite per-stage permutation testing. The sample included both sexes, but sex-stratified analyses were not feasible given the subgroup sizes; sex differences in fNIRS pain responses remain to be tested in larger samples. Age and ethnicity effects could similarly not be assessed, and generalization beyond dental pain remains to be tested.

In conclusion, per-patient calibration with temporal HRF features supports portable fNIRS-based analgesic-state detection under strict subject-independent evaluation, while within-session validation – standard in the field – substantially overestimates performance.

## Data Availability

The data that support the findings of this study are available from the corresponding author upon
reasonable request.

## Acknowledgments

This work was supported by the PI Discretionary Fund within the School of Dentistry/Biologic and Materials Sciences – Prosthodontics, University of Michigan.

## CRediT Author Statement

**Cristian Minoccheri:** Conceptualization, Methodology, Software, Formal analysis, Writing - original draft. **Pangyu Joo:** Data curation. **Xiao-Su Hu:** Data curation, Investigation. **Hafsa Affendi:** Investigation. **Fadi Elayyan:** Investigation. **Angeline Harville:** Investigation. **Alexandre F. DaSilva:** Conceptualization, Methodology, Supervision, Funding acquisition, Writing - review & editing.

## Data and Code Availability

The data that support the findings of this study are available from the corresponding author upon reasonable request. Analysis code for feature extraction, classification, and validation comparison is available at https://github.com/Minoch upon publication.

## Ethical Approval

This study was approved by the University of Michigan Institutional Review Board (protocol number: HUM00164611). All participants provided written informed consent prior to enrollment.

